# Multi-Polygenic prediction of Frailty and its Trajectories highlights Chronic Pain, Rheumatoid Arthritis, and Educational Attainment pathways

**DOI:** 10.1101/2024.05.31.24308260

**Authors:** J P Flint, M Welstead, S R Cox, T C Russ, A Marshall, M Luciano

**Author notes:** Corresponding Author: Jonny Flint, MSc, Lothian Birth Cohorts, Department of Psychology, University of Edinburgh, 7 George Square, Edinburgh EH8 9JZ.

## Abstract

Frailty is a complex ageing-related trait with a growing evidence base for genetic influence. While a single polygenic score (PGS) for frailty has shown predictive value, few studies have examined the joint effect of multiple genetic risks. This study used a multi-polygenic score (MPS) approach to evaluate the combined and relative contributions of 26 PGSs to frailty, measured via the Frailty Index (FI), in two UK cohorts aged 65 and older: the English Longitudinal Study of Ageing (ELSA) and the Lothian Birth Cohort 1936 (LBC1936). Using elastic net regression with repeated cross-validation, we identified chronic pain and depressive symptoms PGSs as the strongest risk predictors of cross-sectional frailty status, while educational attainment, parental longevity, and rheumatoid arthritis PGSs were protective. Compared to single PGS models, MPS models provided improved prediction of frailty levels, explaining up to 4.7% of variance in frailty status – an improvement over the best single PGS (2.5%). To assess whether PGSs also predicted longitudinal frailty progression, we applied generalized additive mixed models (GAMMs) to model age-related trajectories. In ELSA, five PGSs (chronic pain, depressive symptoms, rheumatoid arthritis, educational attainment, and parental death) significantly interacted with age, influencing the rate of frailty change. In LBC1936, consistent though weaker effects were observed for chronic pain and education PGSs. These findings show that polygenic liability shapes both frailty levels and trajectories in later life. Our results support the use of multi-trait genomic models to improve risk prediction and understanding of frailty’s complex aetiology.

## Introduction

Frailty is a multifaceted state associated with ageing and it reflects a decline in physiology and resilience to stressors ^(1)^. Though there is yet to be a unified understanding of what frailty is and how to best measure it, one of the most widely used tools is the frailty index (FI) ^(2,3)^. The FI outlines frailty as a risk index – a cumulative deficit model which totals the number of deficits, such as falls, physical health conditions, grip strength, cognitive functioning, to produce an individualised frailty score – frailty indices often fluctuate with the number of deficits, the criteria suggest a minimum of 30-40 deficits spanning multiple systems, such as health, cognitive, and psychosocial systems related to ageing ^(3,4)^. Higher levels of frailty, reflected by a high frailty score, have been shown to predict adverse outcomes such as hospital stays, disability, and mortality ^(5,6)^.

Other research has shown that frailty is malleable and can be reversed when measured with the FI ^(7,8)^. As the number of people aged over 60 is expected to grow worldwide from 901 million to 1.4 billion from 2015 to 2030 ^(9)^, health conditions like frailty are expected to rise and more people will be living in poorer health in later life ^(10)^. There is, therefore, a necessity to understand how omics, such as genetics, can be used to identify its underlying mechanisms and those at greater risk.

Whilst research investigating the genetic associations of frailty remains in its infancy, the phenotypic associations of frailty have been widely investigated. A systematic review of factors associated with frailty found that the most common factors are mainly sociodemographic factors, such as age, sex, educational level and socioeconomic status ^(11,12)^. Other factors included body weight and level of physical activity, psychological factors such as depressive symptoms, diet quality, neighbourhood, health care quality/access to private healthcare insurance. Furthermore, cross-sectional research investigating phenotypic factors associated with frailty found that loneliness, mobility issues, history of stroke, arthrosis, peripheral vascular disease, cancer, diabetes, hypertension, pain and polypharmacy and drug interactions may predispose an individual to frailty ^(13)^.

Methods which measure the heritable risk of developing a disease via combining the risk of different genetic variants to produce a quantitative score, a polygenic score (PGS), have potential to uncover new information about the molecular underpinnings of this complex multifaceted trait ^(14,15)^. Through data gathered via Genome Wide Association Studies (GWAS), such as GWAS of complex traits like frailty ^(16,17,18)^, PGS can be developed for individuals ^(15)^. Once developed and modelled alongside social and environmental variables, these scores have the potential to be highly predictive for trait and disease prevention – allowing deeper understanding of ageing outcomes and age-related traits and disease. Furthermore, beyond advancing our understanding of the genetic architecture of complex traits, such as frailty, polygenic prediction holds practical risk stratification utility. Identifying individuals at heightened genetic risk for frailty may allow for earlier intervention, even before clinical symptoms manifest. This could inform targeted prevention efforts, such as personalised exercise, nutrition, or social support strategies in midlife or early old age. Moreover, genetic profiles may complement risk modelling around frailty and adverse outcomes, enabling more precise risk stratification in the community.

Our previous work showed that a PGS for frailty can predict frailty, when measured with the FI, at varying ages in later life in the Lothian Birth Cohort 1936 (LBC1936) and the English Longitudinal Study of Ageing (ELSA) ^(19)^. However, if frailty arises from the cumulative effects of its individual components, as the FI states, then just as the FI includes over thirty components to measure frailty, we can approach the genetic prediction of frailty in a similar manner – a multi-polygenic approach ^(20)^. Due to the polygenic and complex nature of frailty, it is informative to aggregate PGSs to investigate their joint predictive power, allowing a more rounded understanding of the genetic contributions to frailty and for refined phenotypic prediction. The GWAS of the FI, in the UK Biobank, tested various PGSs as predictors of FI in univariate linear regression models and found that PGSs for BMI, Inflammatory Bowel Disease (IBD), Waist Hip Ratio (WHR), menarche, grip strength, age at first sexual intercourse, parents’ survival and educational attainment were associated with the FI ^(17)^. The present study improves on this PGS approach by extending it to a multivariate framework using the most recent GWAS to create PGS for multiple variables that are themselves indicators of the FI or expected to relate to them. When modelling genetic variables that are highly correlated with each other, there is risk of multicollinearity ^(21)^. To handle multicollinearity, the PGS are built into a net-elastic regression model to estimate their joint predictive power and order these predictors by their contributing strength to the FI phenotype.

Importantly, this study goes beyond static prediction by investigating whether PGSs also shape the trajectories of frailty across age. Using longitudinal data from ELSA and LBC1936, we model frailty as a nonlinear function of age and test whether genetic risk interacts with age to influence the rate of frailty progression. This allows us to move from assessing whether PGSs are associated with frailty at a given timepoint to whether they predict change in frailty over time – an approach that has particular relevance for understanding ageing processes and informing targeted intervention timing.

Trajectory modelling also helps capture the heterogeneity of ageing patterns across individuals, making the findings more dynamic and clinically informative. To validate the effects found in ELSA, we apply the same longitudinal modelling approach in LBC1936.

## Methods

### Target/Independent Sample

The target sample consisted of the observed genotypes (genome-wide SNPs) and phenotypic data from 5955 adults aged 65-99, mean age 72.73 (SD = 7.2, 2740 males), in ELSA. ELSA is a prospective cohort study which is representative of individuals aged over 50 and living in private households in England ^(22)^. Ethics for ELSA have been approved via the South-Central Berkshire Research Ethics Committee (21/SC/0030, 22nd March 2021). PGS for ELSA were acquired through request to the ELSA genetics team, methods and pre-processing/quality control can be found in the detailed documentation ^(23)^.

An independent test sample was used to validate the findings longitudinally at ∼70, ∼76 and ∼82 years in LBC1936. LBC1936 is an ongoing longitudinal study of older adults living in the community in Edinburgh and surrounding Lothian areas of Scotland, United Kingdom ^(24)^. Individuals were initially recruited based on having been part of the Scottish Mental Survey (1947) and have thus far taken part in 6 waves of testing. Data were drawn from Waves 1, 3, and 5 where appropriate frailty data was collected. At Wave 1 there were 1005 older adults mean age 69.58 (SD = 0.83, n = 1091, 548 males), 697 older adults at Wave 3 mean age 76.30 (SD = 0.68, n = 697, 360 males) and 431 older adults at Wave 5 mean 82.06 (SD = 0.53, n = 431, 209 males). Ethical permission was approved from the Multi-Centre Research Ethics Committee for Scotland (Wave 1: MREC/01/0/56), the Lothian Research Ethics Committee (Wave 1: LREC/2003/2/29), and the Scotland A Research Ethics Committee (Waves 2, 3, 4 and 5: 07/MRE00/58). Written informed consent was obtained from participants at each of the waves. DNA was collected via blood samples from the majority of participants at Wave 1 and genotyping was performed using stringent quality control measures ^(25)^.

### Predictors

As the polygenic scores (PGS) for the ELSA study were either publicly available or obtained upon request from the genetics team, 26 PGSs were identified that either corresponded to traits commonly included in frailty indices or were phenotypically associated with frailty.

While not intended as a direct one-to-one mapping to the items of the Frailty Index (FI), our aim was to capture broad coverage across frailty-relevant domains physical health, cognition, psychosocial factors), akin to how frailty is measured in the literature, using the available PGS from the ELSA repository. To validate findings from ELSA, significant PGSs were then constructed in LBC1936.

Quality control of GWAS summary statistics was conducted using the QCGWAS package ^(26)^, and PGSs in both ELSA and LBC1936 were derived using the same method and software – PRSice-2 ^(27)^. A full list of predictors selected in ELSA and those tested in LBC1936 is provided in the supplementary methods.

### Outcomes

The FI in ELSA was previously constructed from the ELSA dataset ^(28)^. The index consists of 62 deficits, as shown in supplementary Table S1, and a frailty score was created for participants if data were available for 30 out of the 62 possible deficits. Due to skewness in the data, the FI variable was transformed using a square root transformation.

In LBC1936, the FI was previously constructed in the dataset ^(29)^ and contains 30 deficits, including physical, biological, social, psychological, and cognitive deficits, consistent with the FI in ELSA and other research ^(3)^ – the items in the index can be seen in supplementary Table S2. Deficits were either dichotomised as either 0 (absent) or 1 (present); in some cases, 0.5 was used to represent a partially present deficit or were on a continuous scale (such as walking time) on a scale ranging from 0 to 1. For each individual, the number of deficits present was summed and divided by the total number of deficits. Scores ranged from 0 to 1 – with higher scores indicating higher frailty.

Despite there being more deficits in the FI used in ELSA than in LBC1936, guidelines indicate that as long as a minimum of 30 deficits are used to cover the relevant domains (disability, disease, cognitive functioning, psychosocial factors) then differences between number of deficits should not be an issue ^(3)^. Both LBC1936 and ELSA followed the same guidelines when creating the index. For LBC1936 and ELSA FI scores were standardised to allow comparisons when interpreting the results.

### Covariates

Age and sex (the strongest frailty predictors) were controlled in analysis. Four genetic ancestry principal components for LBC1936 and ten ancestry principal components for ELSA were also controlled for in the analyses to account for population stratification – systematic genetic differences due to ancestry differences. Fewer principal components were needed for LBC1936 as it is more genetically homogenous than ELSA. LBC1936 and ELSA only included participants with European ancestry in genotyping.

### Single-polygenic models

To evaluate the benefit of a multi-polygenic approach in predicting frailty, single-polygenic linear regression models, with the 26 PGS, were built in ELSA with the FI as the outcome measure for comparison.

A multiple linear regression model was first fit including sex, age, and ancestry principal components covariates and the outcome (FI) – the null model. Each PGS was then added as a predictor to the null model and re-run (the full model). The variance explained by the PGS (PGS R^2^) is calculated through deducting the R^2^ of the null model from that of the full model for each of the 26 models.

### Multi-polygenic models

The multi-polygenic model was built to test the joint prediction of 26 PGS and rank the prediction of each PGS to the FI outcome. Elastic net regularized regression has been shown to be a useful technique to reduce issues that occur when a magnitude of predictions, such as multiple polygenic scores, lead to overfitting within a traditional multiple linear regression model ^(20)^.

We employed Elastic Net regression to investigate the associations between PGS and the FI in a sample of 5,955 participants for ELSA. Elastic net regularized regression benefits from two regularization techniques – L1 regularization from LASSO regression and L2 regularization from ridge regression through using variable selection and only retaining variables penalizing coefficients for overfitting. Elastic net regression is particularly valuable for PGS prediction with highly correlated genetic signal, as the elastic net regression method will include all highly correlated variables in a grouping effect. The final coefficients in the model allow for the predictors to be ranked by their contribution of prediction to the outcome. The elastic net regression models were computed using the glmnet and caret R package ^(30,31)^. To optimize model performance and prevent overfitting, we used repeated k-fold cross-validation with 10 folds and 10 repetitions. The model explored 10 different values for the tuning parameter (lambda), which balances the penalties of the Elastic Net model. The best-performing model was selected based on minimizing the root mean squared error (RMSE) during cross-validation. We then extracted the optimal lambda and corresponding performance metrics, including RMSE, mean absolute error (MAE), and R-squared (R²). To further validate the stability of the model coefficients, we performed bootstrapping with 1,000 iterations. For each bootstrap sample, the model was refit, and the coefficients were extracted.

With a smaller sample for the LBC1936 cohort and since the Elastic Net model had already been trained and validated on the larger ELSA dataset using repeated k-fold cross-validation we applied the previously-established model, from ELSA, without repeated cross-validation. We explored 10 values of the lambda tuning parameter, and the optimal lambda was selected based on the model with the lowest error. Model coefficients were then extracted for the best-performing lambda. As in the ELSA dataset, we employed bootstrapping with 1,000 resamples to ensure robust coefficient estimation. This approach allowed us to apply a pre-validated model efficiently while ensuring that the coefficients were stable for further validation.

### Longitudinal trajectory models

To assess whether PGSs also predicted changes in frailty over time, we fitted Generalised Additive Mixed Models (GAMMs) using the gamm4 R package. GAMMs are particularly suitable for modelling age-related change as they flexibly accommodate nonlinear patterns while accounting for individual level variation, and have been effectively applied in life-course and ageing research ^(32,33)^. Frailty index scores were modelled as a nonlinear function of age using penalised splines (s(age)) to flexibly capture the curvilinear progression of frailty across older adulthood, which is known to accelerate with age rather than follow a constant, linear increase ^(4,1)^.

This is particularly important in cohort studies, where age-related change may appear nonlinear or attenuated due to dropout and survivor effects. Although frailty generally worsens over time, the trajectories can flatten at older ages because frailer individuals are more likely to drop out or die, leaving a healthier group in later waves. This selective retention may lead to an underestimation of frailty progression in advanced age ^(29)^.

Penalised splines enable this age-dependent acceleration to be captured without imposing rigid assumptions about the shape of change. This spline approach allows for biologically plausible and data-driven age trends without imposing linearity. Interaction smooths (s(age, by = PGS)) were included to test whether genetic liability not only influenced baseline frailty levels but also altered the shape or rate of frailty progression across age. Random intercepts for participant ID were included to account for repeated measures. Participants with fewer than three frailty observations were excluded to ensure reliable estimation of within-person change and to avoid overfitting the spline terms on sparse data. This threshold yielded downstream samples of 5,588 participants in ELSA and 660 participants in LBC1936.

## Results

The mean scores for frailty and age in ELSA and LBC1936 Waves 1, 3, and 5 are shown in Table 1. As expected, mean frailty scores increased with age. Table 1 also reports pairwise correlations of frailty across LBC1936 waves, confirming that frailty is relatively stable over time. Correlations were at minimum moderate (r > 0.5) and mostly strong (r > 0.7), indicating stable individual differences in frailty levels. Correlations between polygenic scores (PGSs) are presented in Supplementary Figures S1 and S2, which display only significant associations (p < 0.05). In ELSA, the most notable correlations were between phenotypically related traits – for example, waist circumference and BMI (r = 0.52), waist circumference and waist-hip ratio (r = 0.54), and educational attainment and general cognitive function (r = 0.40). The PGS for chronic pain also showed a moderate correlation with the frailty PGS (r = 0.33). While some predictors share variance, the magnitude of correlations suggests that multicollinearity is not severe. In LBC1936 (Supplementary Figure S2), the number of PGSs is lower, as only those identified as predictive in ELSA were included. The highest correlation observed was between waist circumference and waist-hip ratio (r = 0.65). Overall, while some PGSs share underlying genetic components, the retained predictors contribute distinct information, justifying their inclusion in the MPS model. To limit redundancy, only one PGS per domain was included where possible, and none exceeded a correlation of r = 0.7.

**Table 1.** Descriptive statistics for age and FI in ELSA and LBC1936 (raw scores), alongside Pairwise correlations of the FI across Wave 1, 3 and 5 in LBC1936.

|  | N | Mean | SD | Range | LBC1936 Wave 1 | LBC1936 Wave 3 | LBC1936 Wave 5 |
| --- | --- | --- | --- | --- | --- | --- | --- |
| **ELSA** |  |  |  |  |  |  |  |
| FI | 5955 | .17 | 0.11 | 0 – 0.68 |  |  |  |
| Age | 5955 | 72.73 | 7.2 | 65 – 99 |  |  |  |
| **LBC1936 Wave 1** |  |  |  |  |  |  |  |
| FI | 1005 | .16 | 0.09 | 0 – 0.49 | 1*** |  |  |
| Age | 1005 | 69.58 | 0.83 | 67.66 – 71.35 |  |  |  |
| **LBC1936 Wave 3** |  |  |  |  |  |  |  |
| FI | 697 | .20 | 0.09 | 0.02 – 0.65 | .71*** | 1*** |  |
| Age | 697 | 76.30 | 0.68 | 74.64 – 77.75 |  |  |  |
| **LBC1936 Wave 5** |  |  |  |  |  |  |  |
| FI | 431 | 0.21 | 0.09 | 0.03 – 0.58 | .57*** | .74*** | 1*** |
| Age | 431 | 82.06 | 0.54 | 80.98-83.19 |  |  |  |

P < 0.05 is flagged with one star (\*), P < 0.01 is flagged with 2 stars (\*\*), P < 0.001 is flagged with three stars (\*\*\*).

### Single models – ELSA

Out of the 26-single score GWAS-PGS models, 18 had significant associations with the FI with the best single-score model coming from the 2019 GWAS-PGS of chronic pain – predicting 2.5% of the variance in the FI (β = - 0.16, 95%CI, 0.13 - 0.18), false discovery rate (FDR) p < 0.001. The PGS on the right of Figure 1 are those that indicate risk to frailty. For example, those who have genetic predisposition to experience Chronic pain have an increased risk of frailty. Whereas the PGS on the left of Figure 1 are those with protective mechanisms. For example, genetic predisposition to having Parental life length (parents living longer) is associated with a decreased risk of frailty.

**Figure 1.**
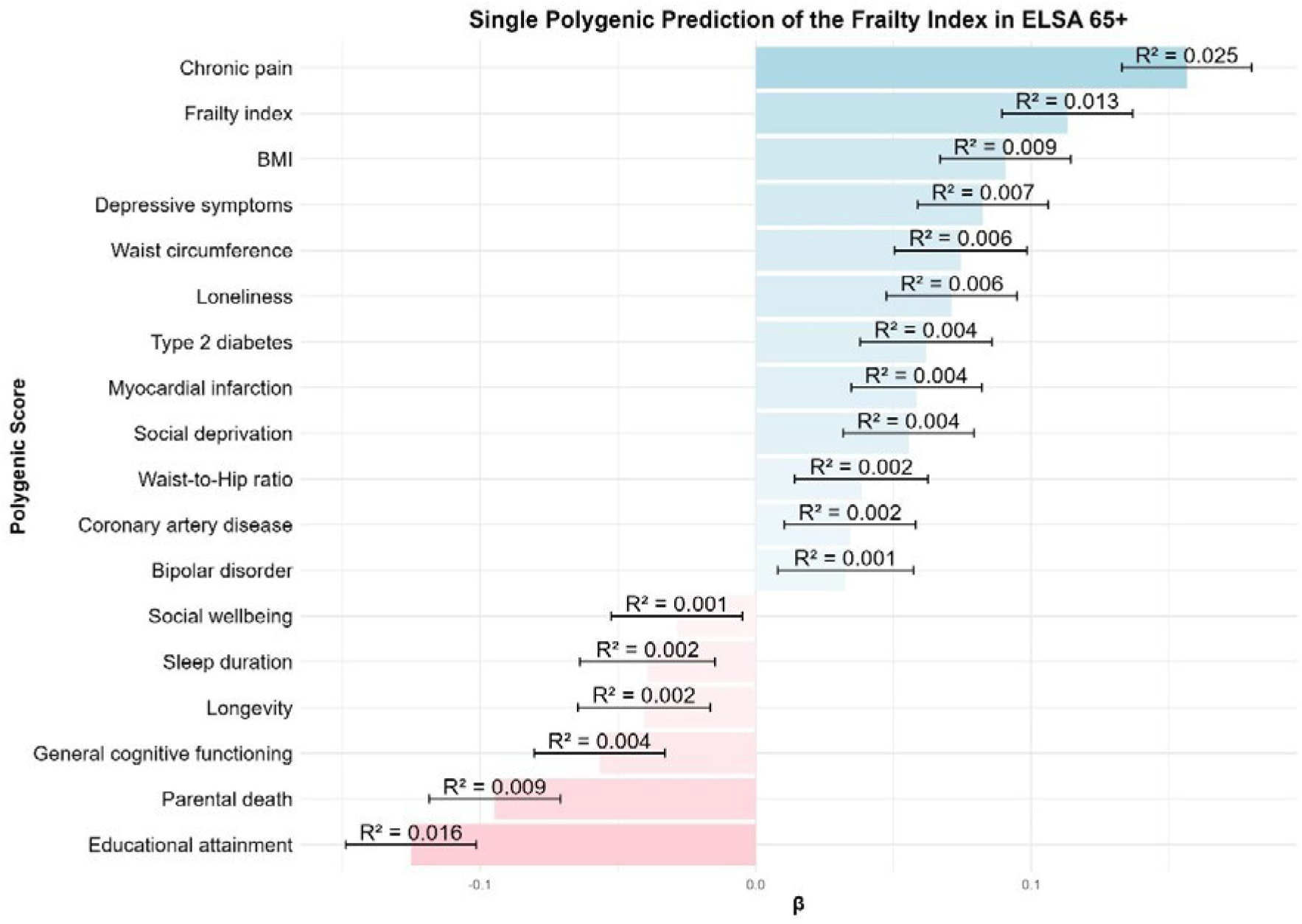
Bar plot of the significant single model PGS with the Frailty Index as the outcome, the y-axis represents the polygenic scores and the x-axis represents beta coefficients with 95% confidence intervals. The R^2^ of each model is displayed on the bars.

### Multi-polygenic models (MPS)

To utilise the benefits of elastic net regression, that is selecting a subset of predictors that are correlated but not redundant to prediction of an outcome, all 26 PGS were modelled into an elastic net regression model with multiple polygenic scores – MPS model. Figure 2 shows the PGS predictors, with covariates included, ranked in effect with standardized coefficients – with those on the right being PGS which exacerbate/worsen frailty and those on the left PGS which protect against frailty. Much like multiple regression models, standardized coefficients represent the contribution of a standard deviation increase in the predictor, in this case PGS, to change in the outcome, in this case the FI, when all other variables in the model have been adjusted for.

**Figure 2.**
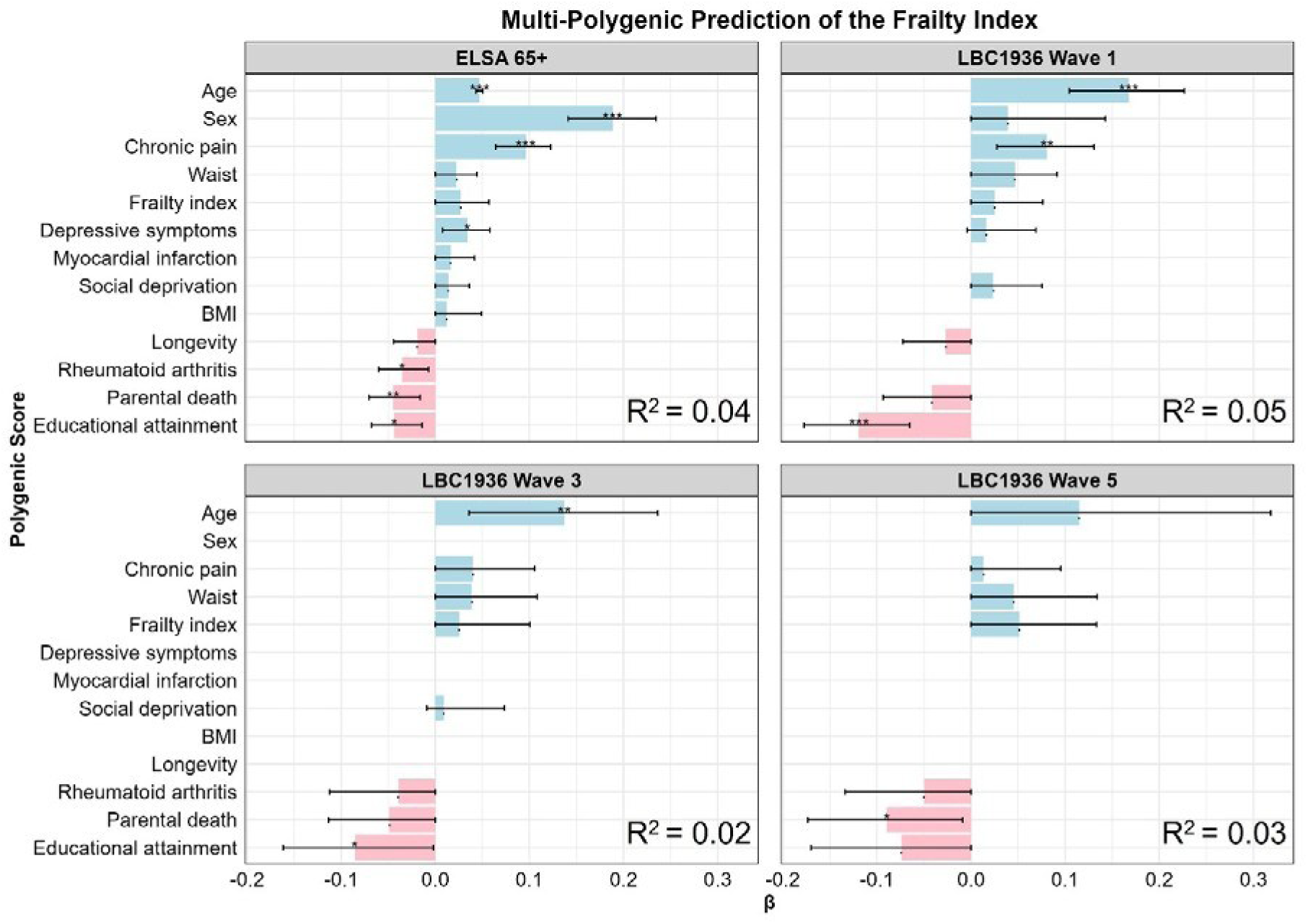
Multi-polygenic prediction of the Frailty Index in ELSA (top left) and LBC1936 (top right: Wave 1; bottom left: Wave 3; bottom right: Wave 5). Bar plots show standardized beta coefficients from Elastic Net regularised regression models including selected polygenic scores (PGSs) and covariates (age, sex, and ancestry principal components). Stars indicate significance after FDR correction: p < 0.05 (*), p < 0.01 (**), p < 0.001 (***). The R² values represent the additional variance in frailty explained by the multi-PGS predictor set beyond covariates alone. Note: Although "Age" and "Sex" are displayed on the y-axis alongside PGSs, they are covariates in the model and not polygenic scores – displayed to show relativity in effect size.

Out of 26 PGS inputted into the model in ELSA, 11 came out as the most relevant predictors, the coefficients for which had not been shrunk to 0. The first plot in Figure 2 shows the standardised beta values and the variance explained through each prediction model. Out of 11 PGS all were significant predictors in the single score models apart from PGS for Rheumatoid arthritis in the single-score models. Interestingly, Rheumatoid Arthritis emerged as a protective PGS for the FI (β = - 0.036, 95%, -0.007 --0.06). The other protective PGS were: Educational attainment (β = - 0.044, 95%, -0.014 - -0.07), Parental death (β = - 0.045, 95%, -0.07 - -0.016), and Longevity (β = -0.020, 95%, -0.04 - 0). On the other side, the strongest predictor of risk of frailty in the MPS model was Chronic pain (β = 0.10, 95%, 0.06 - 0.12).

Alongside PGS for the FI itself (β = 0.027, 95% 0 - 0.057), Depressive symptoms (β = 0.034, 0.007 - 0.058), Waist circumference (β = 0.022, 95%, 0 - 0.04), Myocardial infarction (β = 0.016, 95% 0 - 0.042), BMI (0.012, 95%, 0 - 0.05), Waist circumference (β = 0.022, 95%, 0 - 0.04), and Social deprivation (β = 0.014, 95%, 0 -0.04) were PGS which had risk effects on the FI outcome. Only PGS for Rheumatoid arthritis, Educational attainment, Parental death, Chronic pain and Depressive symptoms were significant upon false discovery rate (FDR) corrections.

### LBC1936 Results (Waves 1, 3, and 5)

The most predictive PGSs identified in ELSA were applied to LBC1936 at Waves 1 (∼age 70), 3 (∼76), and 5 (∼82); results are shown in the third and fourth panels of Figure 2. Across all waves, Educational attainment and Parental death PGSs were consistently associated with lower frailty scores (protective), with educational attainment showing the strongest effects (β range = –0.07 to –0.12), surviving FDR correction at Waves 1 and 3. Additional protective effects were observed for Longevity (Wave 1) and rheumatoid arthritis (Waves 3 and 5), though these did not remain significant after correction.

PGSs for chronic pain, frailty, and waist circumference were the most consistently associated with increased frailty (β range = 0.04 to 0.08), with Chronic pain surviving FDR correction at Wave 1. Social deprivation and Depressive symptoms PGSs showed weaker or marginal associations. At Wave 5, only the Parental death PGS remained significant following correction.

### Longitudinal trajectory models

As shown in Figure 3, in ELSA, PGS showed significant age interaction for Chronic pain (F(2.00) = 52.0, p < .005), Educational attainment (F(3.05) = 7.9, p < .005), Depressive symptoms (F(4.44) = 5.0, p < .005), Rheumatoid arthritis (F(2.64) = 9.7, p < .005), and Parental death (F(4.44) = 11.9, p < .005). The Chronic pain PGS was associated with accelerated frailty progression across age, whereas PGS for Educational attainment, Rheumatoid arthritis, and Parental longevity were associated with more stable or flatter frailty trajectories. The Depressive symptoms score, in contrast, was linked to steeper age-related increases in frailty. These results indicate that multiple domains of polygenic liability – spanning physical, psychological, and socioeconomic dimensions – contribute to variation in how frailty accumulates over time in later life.

**Figure 3.**
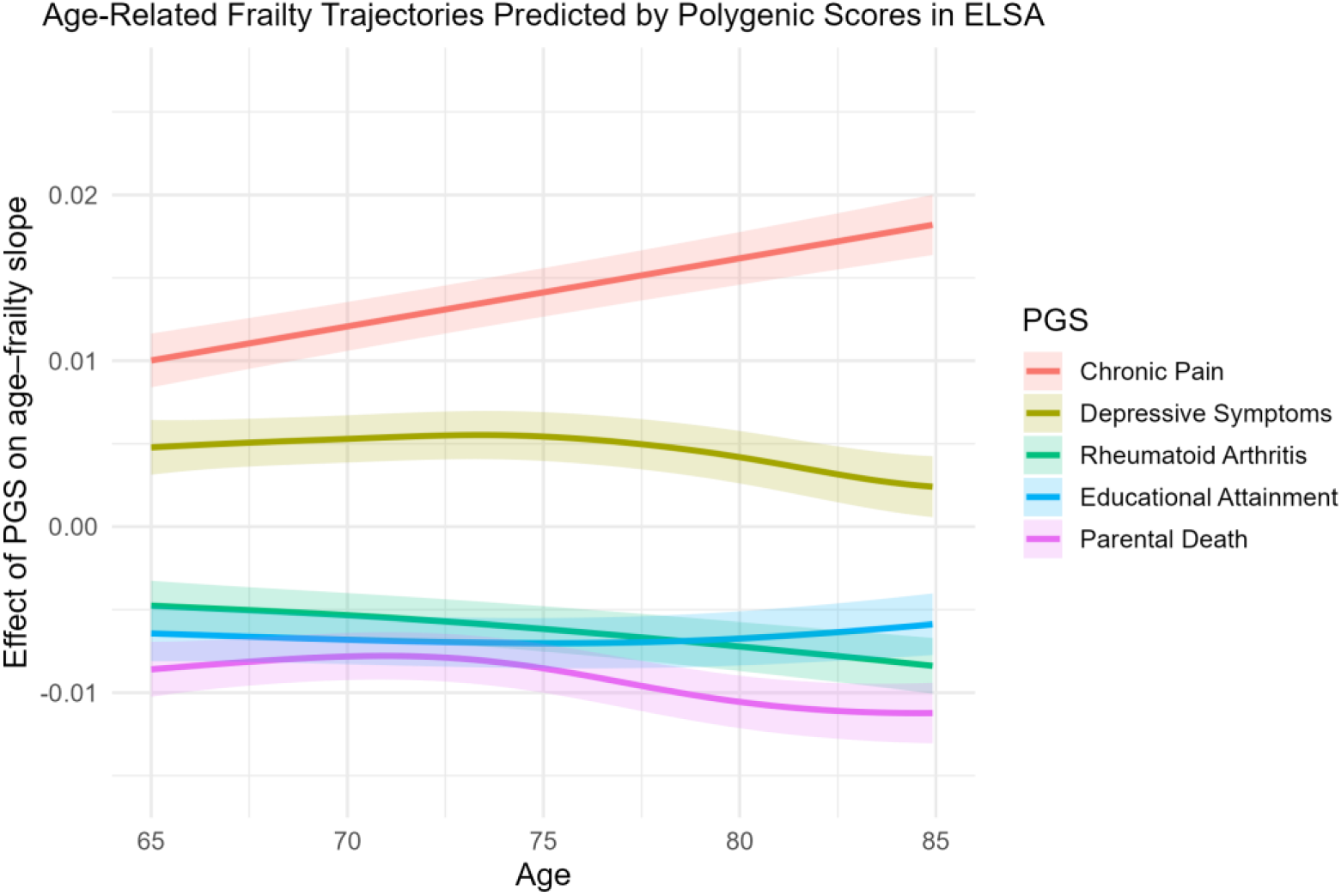
Smooth trajectories from Generalised Additive Mixed Models (GAMMs) showing interaction effects between polygenic scores (PGSs) and age on frailty progression in ELSA. Models include penalised splines for age (s(age)) and interaction smooths (s(age, by = PGS)) to assess whether genetic liability modifies the rate of frailty increase with age. Plots display the estimated smooth effect of each PGS on the age–frailty slope, with 95% confidence intervals. Covariates include age, sex, and ten ancestry principal components. Only PGSs showing significant age interactions are plotted.

To test whether these findings replicated in an independent cohort, we applied the same GAMM approach in the LBC1936 study, focusing on the significant polygenic scores ELSA: Chronic pain and Educational attainment. While the effect sizes in LBC1936 were smaller, both PGS showed statistically significant interactions with age that mirrored the direction of effect observed in ELSA. Specifically, the Chronic pain PGS was associated with faster frailty increases across age (F(2.00) = 3.4, p < .05), while the Educational attainment score predicted a slower rate of frailty accumulation (F(2.00) = 7.3, p < .001). These consistent effects across two independent cohorts strengthen the evidence that genetic liability to Chronic pain and lower Educational attainment contributes to frailty progression during older adulthood. A direct comparison of these effects in ELSA and LBC1936 is shown in Supplementary Figure S3.

## Discussion

By using a multivariate polygenic score approach to predict frailty in British individuals aged 65-99 years, we found robust associations for Chronic pain and Educational attainment at single timepoints and over time. Other PGSs that demonstrated significant associations with the frailty index included Frailty, Waist Circumference, and Socioeconomic deprivation (increasing risk), and Educational attainment, Parental longevity, and Rheumatoid arthritis (decreasing risk). When predicting frailty trajectories in the English Study of Ageing, PGSs for Chronic pain, Educational attainment, Parental longevity, Rheumatoid arthritis, and Deprivation significantly interacted with age, demonstrating divergent influences on the slope of frailty progression. For example, the Chronic pain PGS was associated with steeper frailty increases over time, while the Education attainment PGS buffered age-related frailty growth. These effects were visualized using smooth terms and indicate that PGS not only influences frailty levels cross-sectionally but also the rate of change. In LBC1936, only Chronic pain and Education PGSs showed significant age interactions. Though the magnitude was smaller, the direction of effect was consistent with ELSA. We interpret these differences as reflecting lower variability in frailty, narrower age ranges, and healthier sampling in LBC1936.

In reference to the particular polygenic findings, one may have expected that the Frailty PGS would be the most powerful predictor across the datasets – as this PGS is based on items within the FI itself (outcome measure); however, this was not the case. Chronic pain was the most predictive PGS in the ELSA datasets and at Wave 3 in LBC1936 – with the effect size for PGS Chronic pain almost four times that of the PGS for frailty in ELSA. Chronic pain emerged as the most predictive PGS in both ELSA datasets and LBC1936 datasets. This finding highlights two key points about the measure of frailty. First, frailty is multifaceted: frailty is not a uniform condition; it varies across populations, ages, and cultures. The FI is influenced by the health profile of the population being studied, including how individuals interact with the healthcare system and the priorities of the healthcare system. The weaker predictive power of the FI PGS, when compared to other PGS, raises questions about its construction and how we conceptualise frailty. The FI that was used to build the FI PGS was derived from the UK Biobank. This FI-PGS reflects the health and healthcare interactions of its population. Second, the frailty indices differ in their items, but not criteria, capturing overall functionality or a "health odometer" as individuals age. This flexibility allows the use of readily available data but also presents challenges in measuring frailty as a single aggregated score. For instance, an individual may be ‘cognitively’ frail but still physically mobile, leading to different effects and potentially obscuring important subtypes of frailty. This complexity has been noted in studies modelling phenotypic elements of frailty ^(34)^, which stress the importance of understanding frailty as a dynamic health status that evolves with age, rather than as a static single phenomenon. Thus, the finding that the Chronic pain PGS is more predictive than the FI-PGS may be explained through looking at the genetic architecture of frailty not as a single phenomenon but as a health status that changes across time – meaning the prevalence of the phenomena that make frailty change as a person ages.

When attempting to understand the epidemiology between frailty and pain, our findings can be supported by research that has found some shared mechanisms between the two, suggesting that pain may predispose an individual to developing frailty ^(35)^. Inflammatory markers were shown to be important mediating factors in the progression of frailty and during chronic pain. One example is an abnormal increase in oxidative stress which is present in both pain and the ageing process ^(35)^. The investigation into chronic pain and frailty may also have clinical applications. For instance, a pain treatment plan has been shown to reverse adverse changes in the immune system and reverse neuro-inflammation and restore pro and anti-inflammatory cytokine balancing in the body ^(36)^. Such a plan might be effective in treating and reversing frailty development. However, when multimorbidity and polypharmacy are present pain treatment (medication treatment) might even worsen frailty ^(37,38,39)^.

Despite the clear clinical relevance into the pain-frailty dyad, more research is needed to understand the shared aetiology and direction of influence between the two phenotypes, including any interactions with medication.

The use of elastic net regularization is particularly relevant to disentangling the chronic pain finding and account for multicollinearity – a common issue when modelling multiple PGSs that may be correlated. Elastic net performs grouping, meaning correlated predictors (such as chronic pain and frailty PGSs) can both be retained if they share predictive variance.

Therefore, the coefficients reflect both shared and unique variance across PGSs. While we identified chronic pain as a stronger predictor of frailty than the frailty PGS itself, this may reflect overlapping genetic architecture between these traits. Bidirectional Mendelian randomisation studies have demonstrated a two-way causal relationship between frailty and pain, including both chronic and site-specific pain phenotypes ^(40,41)^. In addition, a twin study from TwinsUK found substantial shared genetic aetiology between frailty and chronic widespread pain ^(42)^, supporting the notion of common underlying biological pathways. These findings reinforce the value of joint polygenic modelling when investigating complex, multifactorial traits like frailty.

Various PGS were shown to have protective associations with the FI across the datasets/waves. In regard to the PGS for Educational attainment, in a similar study to ELSA – the Health and Retirement Study, higher PGS for Educational attainment was associated with lower frailty levels, when measured with the FI ^(43)^. However, the same study found the effects of PGS on educational attainment were not present in participants older than 80 years – our study contradicts these findings as the effect for the Educational attainment PGS was still present in LBC1936 Wave 5 (age ∼82). Nevertheless, our study did find that the association weakened with advancing age, similar to the Health and Retirement Study findings. It is important to note that genetics, to a degree, influences educational attainment, partly via the personal characteristics that help individuals achieve academically ^(44)^. Thus, when we discuss genetic variance of educational attainment, we must reflect on these heritable traits/characteristics, including cognitive traits and personality traits such as conscientiousness that also relate to healthy life styles, money for health care – mainly driven via the association between educational attainment and socioeconomic level ^(44,45, 46)^. The PGS for Educational attainment is different to the other PGS, as many of the others are direct indicators of the FI. There are applications for social researchers/statisticians here to further understand the relationship between society, educational attainment PGS, and outcomes like frailty.

Besides the PGS for Educational attainment, PGS for Parental death was also found to be protective against frailty. Previous research has shown that protective mechanisms have been found between a parent living a long life and physical functioning – a phenotype correlated with frailty ^(45,46,47)^. Unlike PGS for Educational attainment and the lifespan of a parent, PGS for Rheumatoid arthritis and its relationship with frailty is not well studied or understood. The finding that PGS for Rheumatoid arthritis is a protective PGS for frailty is surprising – as one would expect the inflammation pathways associated with rheumatoid arthritis would put one at risk to frailty. A possible mediation pathway could be the role of anti-inflammatory drugs that people with rheumatoid arthritis may be taking. Some evidence has demonstrated that anti-inflammatory medications may lower levels of inflammatory biomarkers ^(48)^ – such medications could also lower the risk of frailty. However, there is mixed data on the association between anti-inflammatory medications and frailty. For example, long-term aspirin use was associated with a 15% lower risk of frailty ^(49)^. However, when extending this work the researchers found long-term use with other Nonsteroidal anti-inflammatory agent (NSAID) medication was associated with an increased prevalence of frailty, even after consideration of multimorbidity and health behaviours ^(50)^. Further research around rheumatoid arthritis, medication use, and frailty is an important avenue of research that is stimulated by the findings of this paper.

The findings from this study could also highlight the issue with the measure of frailty at different ages and biases in sampling. Many of the GWAS come from UK Biobank samples which include younger/middle aged adults who are more likely to be healthy, have higher educational attainment and fewer health conditions – the healthy participation bias ^(51)^. ELSA and LBC1936 may also suffer from the healthy volunteer effect ^(52)^, alongside those who remain in the sample being more healthy – healthy survivor effect. The differences between these cohorts are also important as ELSA is representative of the UK population through its sampling of England ^(22)^. However, LBC1936 is unique to an area in Lothian, Scotland, and is less representative of the Scottish population with respect to cognitive functioning and mortality/longevity.

Despite the shortcomings, the current study has several strengths. The analysis first being carried out in ELSA and validated longitudinally in LBC1936 increased the validity and generalisability of the findings – helping question and validate any spurious associations. The method also dealt with multicollinearity and overfitting by using Elastic net regularised regression – a method which has been shown to be useful when fitting multiple genetic instruments ^(20)^. Future research might usefully explore the polygenic nature of frailty but with different samples to address the differing strengths of PGS. In particular, replication of our effects in countries other than England and Scotland is needed to understand several mixed findings and direction of PGS effects between ELSA and LBC1936 (for example the PGS for Depression). It may be that interactions with environmental and social factors (such as differences in health care service, life adversity) are masking or exacerbating the genetic effect. It may also be valuable to model frailty with different outcomes, such as the Fried Phenotype and clinical frailty measures.

In conclusion, our results demonstrate that frailty is a genetically complex, multidomain trait best understood through multi-PGS frameworks. We identified novel relationships between multiple PGSs and frailty, measured both cross-sectionally in older age and longitudinally over time. Chronic pain and Educational attainment consistently emerged as core PGS predictors, influencing both frailty levels and trajectories. However, it is important to emphasise that-even in combination- these PGSs explained a relatively modest proportion of variance (up to ∼5%), underscoring their current limitations for individual-level risk prediction. These findings are more informative at the group or population level and should be interpreted as reflecting general liability rather than deterministic outcomes. For Chronic pain, predictive utility may reflect shared biological mechanisms relevant to frailty onset, while the Educational attainment PGS – a trait established decades before frailty develops – may capture cognitive, behavioural, or environmental pathways linked to healthier ageing. As ageing research advances, integrating genetic measures across cognitive, physical, and psychosocial domains will be crucial for identifying at-risk subgroups and tailoring intervention windows. Bridging genomic insights with interdisciplinary perspectives ensures that frailty development is understood as not just a biological construct but also a social and behavioural construct.

## Data Availability

Availability of data and material: Data was obtained from the Lothian Birth Cohort 1936, more information can be found at https://lothian-birth-cohorts.ed.ac.uk/ . The English Study of Ageing data: ukdataservice.ac.uk/datacatalogue//series/series?id=200011. Any further data not found in such sources are available on request.

## Author Contributions

Concept and Design J Flint, M Luciano; Data analysis: J Flint; Drafting of the manuscript: J Flint; Critical revision of the manuscript: J Flint, M Luciano, T C Russ, A Marshall, SR Cox; Administrative technical or material support: M Welstead, C Eke

The LBC1936 is supported by the Biotechnology and Biological Sciences Research Council, and the Economic and Social Research Council [BB/W008793/1], Age UK (The Disconnected Mind Project, which also supported MW), the Milton Damerel Trust, and The University of Edinburgh. SRC is supported by a Sir Henry Dale Fellowship jointly funded by the Wellcome Trust and the Royal Society (221890/Z/20/Z). ELSA is funded by the National Institute on Aging (R01AG017644), and by UK Government Departments coordinated by the National Institute for Health and Care Research (NIHR). No editorial service was provided.

## Funding Acknowledgements

This research was funded by the Legal & General Group (research grant to establish the independent Advanced Care Research Centre at the University of Edinburgh). The funder had no role in the conduct of the study, interpretation, or the decision to submit for publication. The views expressed are those of the authors and not necessarily those of Legal & General.

## Availability of data and material

Data was obtained from the Lothian Birth Cohort 1936, more information can be found at https://lothian-birth-cohorts.ed.ac.uk/. The English Longitudinal Study of Ageing data: https://ukdataservice.ac.uk/. Any further data not found in such sources are available on request.

