## Supplementary materials for "Multi-Polygenic prediction of Frailty and its Trajectories highlights Chronic Pain, Rheumatoid Arthritis, and Educational Attainment pathways"

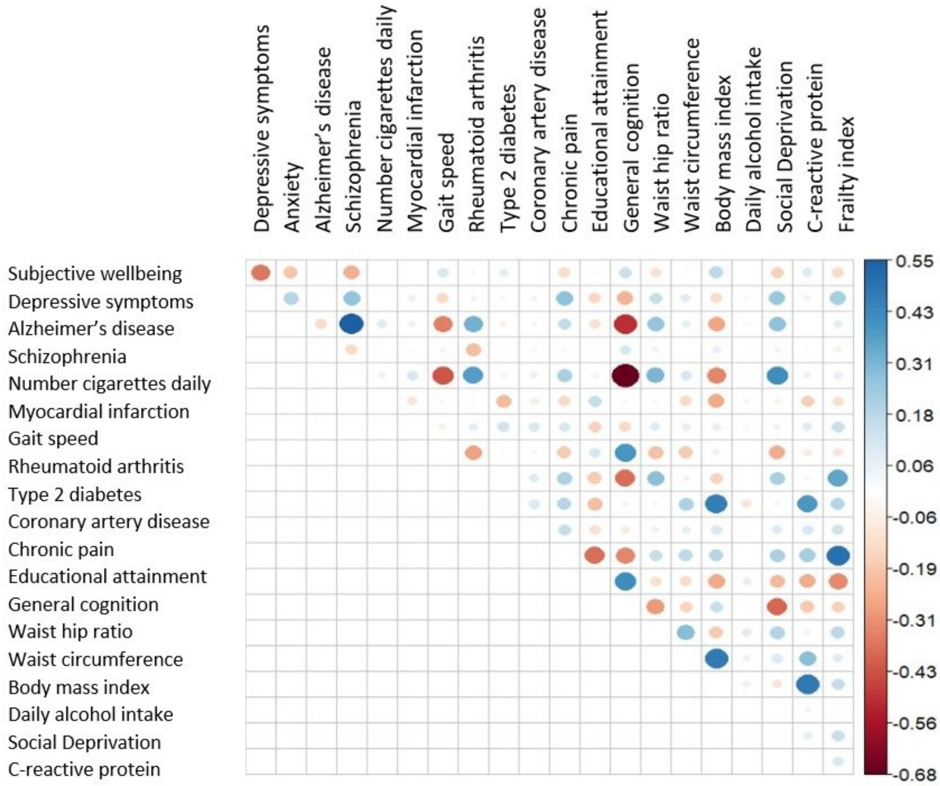


Supplementary Figure S1. ELSA correlation matrix for 26 PGS, presented are any significant correlations ($p < 0.05$).

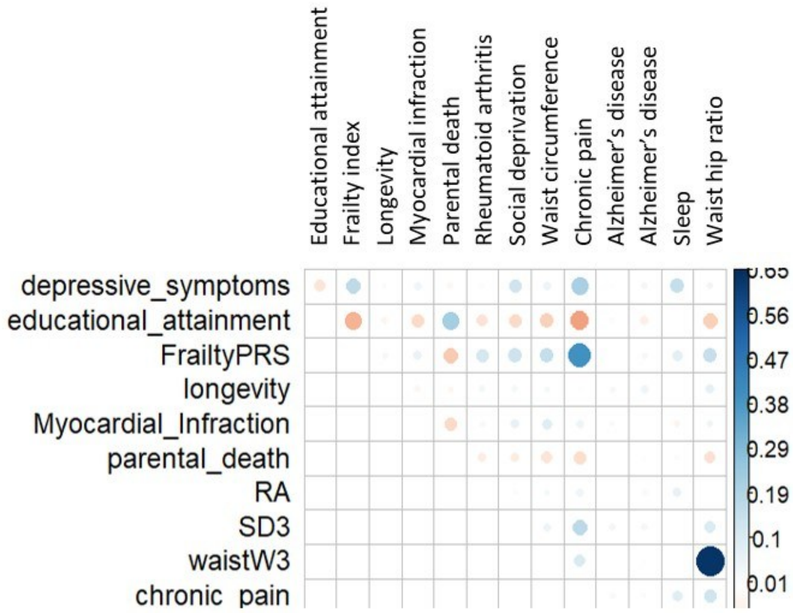


Supplementary Figure S2. LBC1936 (full sample – Wave 1) Correlation Matrix for predictive PGS, presented are any significant correlations ($p < 0.05$).

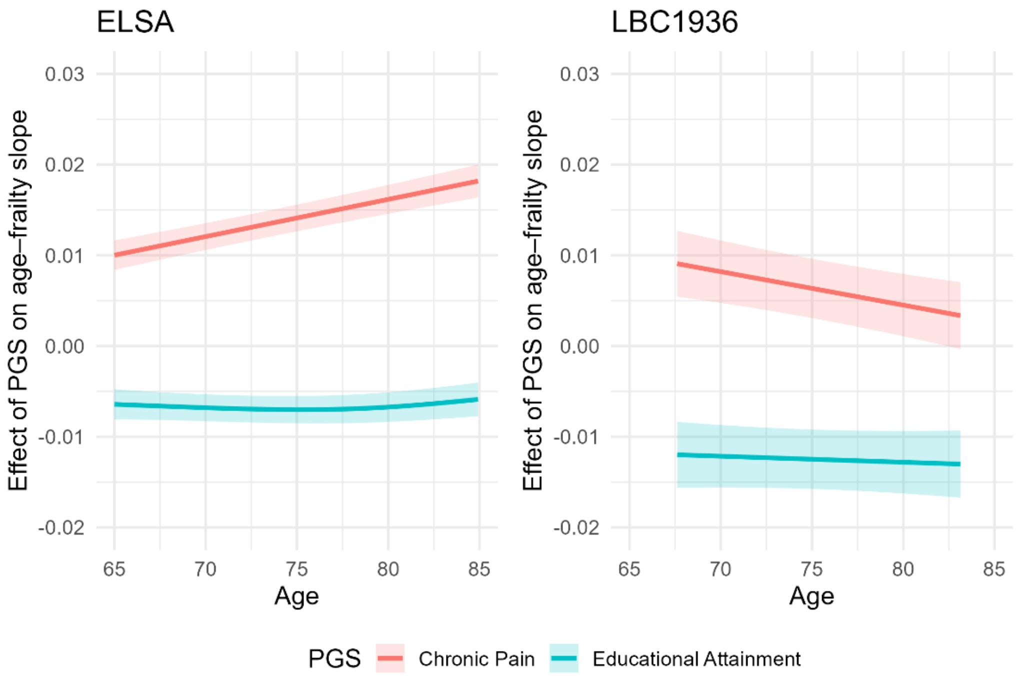


Supplementary Figure S3. ELSA and LBC1936 comparison of the trajectories from Generalized Additive Mixed Models (GAMMs) showing interaction effects between chronic pain, educational attainment and age on frailty progression.

Supplementary Figure S2. LBC1936 (full sample – Wave 1) Correlation Matrix for predictive PGS, presented are any significant correlations ($p < 0.05$).

### Supplementary Table ST1: Elements comprising the frailty index in LBC1936

| Items | Coding | Cut offs based on |
|---|---|---|
| Systolic Blood Pressure | <5$^{th}$ percentile (1), 5$^{th}$-20$^{th}$ percentile (0.5), >20$^{th}$ percentile (0) | Recommended technique (Theou et al., 2015) |
| Diabetes | Yes (1) or No (0) | Already binary variable |
| High Cholesterol | Yes (1) or No (0) | Already binary variable |
| Heart problem | Yes (1) or No (0) | Already binary variable |
| Stroke or mini stroke | Yes (1) or No (0) | Already binary variable |
| Crampy pains in calves | Yes (1) or No (0) | Already binary variable |
| Problems with blood circulation | Yes (1) or No (0) | Already binary variable |
| Thyroid Disorder | Yes (1) or No (0) | Already binary variable |
| Cancer | Yes (1) or No (0) | Already binary variable |
| Parkinson's disease | Yes (1) or No (0) | Already binary variable |
| Dementia | Yes (1) or No (0) | Already binary variable |
| Arthritis | Yes (1) or No (0) | Already binary variable |
| Any other chronic disease | Yes (1) or No (0) | Already binary variable |
| Polypharmacy | >4 medications (1), ≤4 medications (0) | Recommended technique (Theou et al., 2013) |
| Body Mass Index | 18.5 to <25 (0), 25 to <30 (0.5), <18.5 or >equal to 30 (1) | Recommended technique (Chamberlain, Sauver, et al., 2016) |
| 6m walk time (gait speed) | >10 seconds or physically unable (1), <10 seconds (0) | Recommended technique (Hoogendijk et al., 2017) |

| Items | Coding | Cut offs based on |
|---|---|---|
| Able to stand up from a chair | Yes (1) or No (0) | Already binary variable |
| Grip strength (strongest hand and stratified by sex and BMI) | <5th percentile (1), 5th-20th percentile (0.5), >20th percentile (0) | Recommended technique (Theou et al., 2015) |
| Townsend Disability Scale | 11 – 18 (1), 0 -10 (0) | Recommended technique (Fiona Elaine Matthews et al., 2016) |
| Peak Expiratory Flow rate (stratified by sex) | <5th percentile (1), 5th-20th percentile (0.5), >20th percentile (0) | Recommended technique (Theou et al., 2015) |
| Forced expiratory volume (stratified by sex) | <5th percentile (1), 5th-20th percentile (0.5), >20th percentile (0) | Recommended technique (Theou et al., 2015) |
| Depression | 11 -21 (1), 8 – 10 (0.5), 0 – 7 (0) | Recommended technique (Zigmond & Snaith, 1983) |
| Anxiety | 11 -21 (1), 8 – 10 (0.5), 0 – 7 (0) | Recommended technique (Zigmond & Snaith, 1983) |
| MMSE | <10 (1), 11-17 (0.75), 18 – 20 (0.5), 20 – 24 (0.25), >24 (0) | Recommended technique (Searle et al., 2008) |
| Digit Symbol | <5th percentile (1), 5th-20th percentile (0.5), >20th percentile (0) | Recommended technique (Theou et al., 2015) |
| Block Design | <5th percentile (1), 5th-20th percentile (0.5), >20th percentile (0) | Recommended technique (Theou et al., 2015) |
| Verbal Fluency | <5th percentile (1), 5th-20th percentile (0.5), >20th percentile (0) | Recommended technique (Theou et al., 2015) |
| Matrix Reasoning | <5th percentile (1), 5th-20th percentile (0.5), >20th percentile (0) | Recommended technique (Theou et al., 2015) |
| Reaction time test | <5th percentile (1), 5th-20th percentile (0.5), >20th percentile (0) | Recommended technique (Theou et al., 2015) |
| Delayed recall | <5th percentile (1), 5th-20th percentile (0.5), >20th percentile (0) | Recommended technique (Theou et al., 2015) |

**Supplementary Table ST2: Elements comprising the frailty index score and their seven domains in ELSA**

| Variable name | Domain (taken from elsa questionnaire) | Description |
|---|---|---|
| hemobwa | mobility | Difficulty walking 100m |
| hemobsi | mobility | Difficulty sitting 2 hrs |
| hemobch | mobility | difficulty getting up from chair after sitting long periods |
| hemobcs | mobility | Difficulty climbing several flights of stairs without resting |
| hemobcl | mobility | Difficulty climbing one flight of stairs without resting |
| hemobst | mobility | Difficulty stooping, kneeling or crouching |
| hemobre | mobility | Difficulty reaching or extending arms above shoulder level |
| hemobpu | mobility | Difficulty pulling or pushing large objects |
| hemobli | mobility | Difficulty lifting or carrying weights over 10 pounds (4.54kg) |
| hemobpi | mobility | Difficulty picking up a 5p coin from a table |
| headldr | activities of daily life | Difficulty dressing, including putting on shoes and socks |
| headlwa | activities of daily life | difficulty walking across a room |
| headlba | activities of daily life | Difficulty bathing or showering |
| headlea | activities of daily life | Difficulty eating, such as cutting up food |
| headlbe | activities of daily life | Difficulty getting in and out of bed |
| headlwc | activities of daily life | Difficulty using the toilet including getting up or down |
| headlma | activities of daily life | Difficulty using map to figure out how to get around strange place |
| headlpr | activities of daily life | Difficulty preparing a hot meal |
| headlsh | activities of daily life | Difficulty shopping for groceries |
| headlph | activities of daily life | Difficulty making telephone calls |
| headlme | activities of daily life | Difficulty taking medications |
| headlhg | activities of daily life | Difficulty doing work around the house or garden |
| headlmo | activities of daily life | Difficulty managing money, eg paying bills, keeping track of |

| | | |
|---|---|---|
| | | expenses |
| hedimbp | CV | High bp dx |
| hediman | CV | Angina dx |
| hedimmi | CV | Heart attack |
| hedimhf | CV | Congestive heart failure |
| hedimar | CV | Abnormal heart rhythm |
| hedimdi | CV | Diabetes or high blood sugar |
| hedimst | CV | Stroke dx |
| hediblu | Chronic | Lung disease dx |
| hedibas | Chronic | Asthma dx |
| hedibar | Chronic | Arthritis dx |
| hedibos | Chronic | Osteoporosis |
| hedibca | Chronic | Cancer dx |
| hedibpd | Chronic | Parkinson's dx |
| hedibps | Chronic | Psychiatric condition |
| hedibad | Chronic | Alzheimer's dx |
| hedibde | Chronic | Dementia dx |
| psceda | Psych | Whether felt depressed much of the time during the past week |
| pscedb | Psych | Whether felt everything they did during the past week was an effort |
| pscedc | Psych | Whether felt their sleep was restless during the past week |
| pscedd | Psych | Whether was happy much of the time during the past week /R |
| pscede | Psych | Whether felt lonely much of the time during the past week |
| pscedf | Psych | Whether enjoyed life much of the time during the past week /R |
| pscedg | Psych | Whether felt sad much of the time duing the past week |
| pscedh | Psych | Whether could not get going much of the time during the past week |
| hehelf | General | Self-reported general health |
| heeye | General | Self-reported eyesight (while using lenses if appropriate) |
| hehear | General | Self-reported hearing (while using hearing aid if appropriate) |
| hefla | General | Whether fallen down since last interniew |

| | | |
|---|---|---|
| hefrac | General | Whether has fractured hip |
| heji | General | Whether had joint replacement |
| mmpain | General | Timed walk: whether had pain whilst walking |
| cfdatd | Memory | Whether correct day of month given |
| cfdatm | Memory | Whether correct month given |
| cfdaty | Memory | Whether correct year given |
| cfday | Memory | Whether correct day given |
| cfmem | Memory | Whether prompt given for prospective memory test (remembering to write initials) |
| cflisenq | Memory | Refers to cflisen |
| cfaniq | Memory | Refers to cfani |
| cflisdq | Memory | Refers to cflisd |

Supplementary methods

*The 26 PGS and the sources/summary statistics*

**Coronary Artery disease**

CARDIoGRAM 2,420,360 783,413 Schunkert et al.

(2011)[36] www.cardiogramplusc4d.org (cad.add.160614.website.txt)

**Type II Diabetes**

DIAGRAM 2,473,441 761,488 Morris et al. (2012)[37] http://www.diagram-consortium.org/downloads.html (DIAGRAMv3.2012DEC17.txt).

**General cognitive function**

CHARGE 2,473,946 795,327 Davies et al. (2015)[38] https://www.ncbi.nlm.nih.gov/projects/gap/cgi-bin/study.cgi?study_id=phs000930.v6.p1

**Rheumatoid arthritis**

8,747,962 1,100,616 Okada et al. (2014)[39] http://plaza.umin.ac.jp/~yokada/datasource/software.htm

**Myocardial infarction**

CARDIoGRAM 9,289,491 1,299,282 CARDIoGRAMplusC4D

Consortium. (2015)[40] www.cardiogramplusc4d.org (mi.add.030315.website.txt)

**Longevity**

CHARGE 2,588,525 757,472 Broer et al. (2014)[41] https://grasp.nhlbi.nih.gov/FullResults.aspx

**Sleep Duration**

32,449,020 948,331 Lane et al (2017)[43] http://biobank.ctsu.ox.ac.uk/

**Body Mass Index**

GIANT 2,554,623 795,650 Locke et al. (2015)[45] https://www.broadinstitute.org/collaboration/giant/index.php/GIANT_consortium_data_files

**Waist circumference**

GIANT 2,565,407 801,114 Shungin et al.

(2015)[46]

https://www.broadinstitute.org/collaboration/giant/index.php/GIANT_consortium_data_files;

WC: GIANT 2015 WC COMBINED EUR.txt.gz

**Waist-to-hip ratio**

GIANT 2,542,431 801,207 https://www.broadinstitute.org/collaboration/giant/index.php/GIANT_con

**Educational Attainment**

3 SSCAG 10,101,242 1,325,851 Lee et al. (2018)[28] https://www.thessgac.org/data (https://www.dropbox.com/s/ho58e9jmytmpaf8/GWAS_EA_excl23andMe.txt?dl=0)

**Social Deprivation** - 15,732,391 1,341,112 Hill et al (2016) [29] https://grasp.nhlbi.nih.gov/FullResults.aspx

**Alzheimer's disease** IGAP 7,055,881 1,191,420 Lambert et al. (2013)[30] http://web.pasteur-lille.fr/en/recherche/u744/igap/igap_download.php

**Depressive symptoms** SSCAG 6,524,474 1,187,563 Okbay e al. (2016)[26] https://www.thessgac.org/data

**Anxiety Disorders (factor score)** ANGST 6,306,612 1,137,311 Otowa et al. (2016)[33] https://www.med.unc.edu/pgc/results-and-downloads

**Subjective Well-Being**

SSCAG 2,268,674 748,500 Okbay e al (2016)[26] Provided by request

**Number of cigarettes smoked per day** TAG 2,459,118 803,092 https://www.med.unc.edu/pgc/results-and-downloads (tag.cpd.tbl.gz)

**Daily Alcohol Intake** - 2,462,742 800,524 Schumann et al (2016)[48] https://grasp.nhlbi.nih.gov/FullResults.aspx

**Schizophrenia (2014)**

PGC 9,444,230 1,278,742 Ripke et al. (2014)[35] https://www.med.unc.edu/pgc/results-and-downloads (scz2.snp.results.txt.gz)

**Bipolar disorder**

Purcell, S.M., et al., Common polygenic variation contributes to risk of schizophrenia and bipolar disorder. Nature, 2009. 460(7256): p. 748-52.

**Parental death**

Pilling LC, Kuo CL, Sicinski K, Tamosauskaite J, Kuchel GA, Harries LW, Herd P, Wallace R, Ferrucci L, Melzer D. Human longevity: 25 genetic loci associated in 389,166 UK biobank participants. Aging (Albany NY). 2017 Dec;9(12):2504.

**Gait speed**

Ben-Avraham D, Karasik D, Verghese J, Lunetta KL, Smith JA, Eicher JD, Vered R, Deelen J, Arnold AM, Buchman AS, Tanaka T. The complex genetics of gait speed: genome-wide meta-analysis approach. Aging (Albany NY). 2017 Jan;9(1):209.

**Chronic pain**

Johnston KJ, Adams MJ, Nicholl BI, Ward J, Strawbridge RJ, Ferguson A, McIntosh AM, Bailey ME, Smith DJ. Genome-wide association study of multisite chronic pain in UK Biobank. PLoS genetics. 2019 Jun 13;15(6):e1008164.

**Loneliness**

Gao J, Davis LK, Hart AB, Sanchez-Roige S, Han L, Cacioppo JT, Palmer AA. Genome-wide association study of loneliness demonstrates a role for common variation. Neuropsychopharmacology. 2017 Mar;42(4):811-21.

**CRP**

Said S, Pazoki R, Karhunen V, Võsa U, Ligthart S, Bodinier B, Koskeridis F, Welsh P, Alizadeh BZ, Chasman DI, Sattar N. Genetic analysis of over half a million people characterises C-reactive protein loci. Nature communications. 2022 Apr 22;13(1):2198.

**Frailty Index**

Atkins JL, Jylhävä J, Pedersen NL, Magnusson PK, Lu Y, Wang Y, Hägg S, Melzer D, Williams DM, Pilling LC. A genome-wide association study of the frailty index highlights brain pathways in ageing. Aging Cell. 2021 Sep;20(9):e13459.
